# Sensitivity evaluation of 2019 novel coronavirus (SARS-CoV-2) RT-PCR detection kits and strategy to reduce false negative

**DOI:** 10.1101/2020.04.28.20083956

**Authors:** Yunying Zhou, Fengyan Pei, Li Wang, Huailong Zhao, Huanjie Li, Mingyu Ji, Weihua Yang, Qingxi Wang, Qianqian Zhao, Yunshan Wang

## Abstract

An ongoing outbreak of pneumonia associated with SARS-CoV-2 has now been confirmed globally. In absence of effective vaccines, infection prevention and control through diagnostic testing and quarantine is critical. Early detection and differential diagnosis of respiratory infections increases the chances for successful control of COVID-19 disease. The nucleic acid RT-PCR test is regarded as the current standard for molecular diagnosis with high sensitivity. However, the highest specificity confirmation target ORF1ab gene is considered to be less sensitive than other targets in clinical application. In addition, a large amount of recent evidence indicates that the initial missed diagnosis of asymptomatic patients with SARS-CoV-2 and discharged patients with “re-examination positive” may be due to low viral load, and the ability of rapid mutation of coronavirus also increases the rate of false negative results. We aimed to evaluate the sensitivity of different nucleic acid detection kits so as to make recommendations for the selection of validation kit, and amplify the suspicious result to be reportable positive by means of simple continuous amplification, which is of great significance for the prevention and control of the current epidemic and the discharge criteria of low viral load patients.

## INTRODUCTION

The coronavirus that caused the outbreak was identified in the case of viral pneumonia in Wuhan in 2019[1–3], and was named 2019-nCoV/SARS-CoV-2 by the World Health Organization (WHO) [2, 4, 5]. SARS-CoV-2 belongs to the coronavirus genus β and its genome is single-stranded, non-segmented positive-sense RNA[6], which is the seventh known coronavirus that can infect humans[1, 7]. Similar to other pathogenic RNA viruses, the genetic material RNA is the first marker to be detected. Nucleic acid detection or sequencing is currently used in conjunction with pulmonary CT for clinical diagnosis of COVID-19[8, 9]. As the course of the disease progresses, antibodies IgM and IgG will be produce by the human immune system. Although, antibody tests play a major role in monitoring the response to future immunization strategies and demonstrating previous exposure/immunity, the antibody positive rate often lags behind the nucleic acid detection[10–12], and cross-reactions existed in SARS-CoV antigen with autoantibodies [13].

Theoretically, fluorescence quantitative RT-PCR detection is widely used as the molecular diagnosis standard for SARS-CoV-2[14, 15]. Lately, the analysis showed that the pattern of viral load change in COVID-19 patients was similar to that in patients with influenza, but different from that in SARS and MERS (whose viral load peaked about 10 days after the onset of symptoms) [16–19]. At present, a large number of rapid gene detection technologies have been developed in succession, which has great value for the screening of potential infectors and virus detection. However, with too much emphasis on the “fast” characteristic, it is bound to cause a certain degree of sacrifice in other performances. Due to the lack of validation of clinical samples, SHERLOCK technology based on CRISPR/cas13 cannot be used in the clinical diagnosis of SARS-CoV-2[20]; mNGS (macrogenomic sequencing) also faces the challenges of long detection cycle, complex process [21]; Although LAMP method is very sensitive[21, 22], the low load virus will still lead to false negative or spontaneous negative signals of thermostatic technology.

In COVID-19 patients, RT-PCR detection could be positive as early as one day before the onset of symptoms, while most COVID-19 patients cannot be detected before premorbid because of the low copy number of the virus[2, [7, 17, 23]. In addition, some discharged patients appearing “re-examination positive” situation is also because of the persistence of a small number of viruses. Unfortunately, the positive rate of RT-PCR detection of SARS-CoV-2 is only 30%-50% at present[24, 25] due to improper sample collection, storage, and error detection [26]. Furthermore, once the target gene mutated or deleted, the test results will be invalid [27, 28].

RT-PCR nucleic acid detection not only has a high false negative rate[29], but also has a low sensitivity[30]. Currently, the approved nucleic acid detection kits of the SARS-CoV-2 genome are based on the most conserved and specific open reading frame 1ab (ORF1ab), Envelope protein (E) and nucleocapsid protein (N)[6, 31, 32]. Athough ORF1ab is the highest specificity confirmation target gene, but is considered to be less sensitive than other targets in clinical application[33], so does the pattern of ORF1ab positive reports cause missed tests? Is it feasible to report based on positive N or E genes? Clinically, it is recommended that samples with suspicious results or single channel positive results should be re-examined with another manufacturer’s kit or method. However, what is the basis for choosing the validation kit? This is a problem that needs to be solved.

### Material and Methods

#### Patients

10 confirmed cases of COVID-2019 patients (2 female, 8 male, 5–50 years old) were collected from January to February 2020, in Jinan Central Hospital Affiliated to Shandong University and Jinan Infectious Disease Hospital, Shandong University, which were diagnosed by clinical symptoms, lung CT and nucleic acid test. And 100 suspected cases were collected in the first institution listed above, which had symptoms of fever, dry cough and pneumonia image. This research was approved by the Ethics Commission of Jinan Central Hospital and with informed consent of the patient.

#### Specimen collection

Nasopharyngeal and oropharyngeal swab specimens were collected with synthetic fiber swabs under the guideline of the Chinese Centre for Disease Control and Prevention (China’s CDC) (http://www.chinacdc.cn/ikzt/crb/zl/szkb_11803/jszl_11815/202003/t20200309_214241.html). And the two swabs from nasopharyngeal and oropharyngeal were inserted into one sterile tube containing 3 ml of Virus preservation solution. In addition, environmental specimens were collected from surface in direct contact with the patient, such as inner side of the mask, phone, doorknob, bedside, and etc. Each surface was wiped with one synthetic fiber swab, and then inserted the swab into a sterile tube listed above.

#### Virus RNA extractions

The virus RNA was extracted using magnetic bead method strictly according to the instructions of Nucleic acid extraction kit (Shanghai Zhijiang Biotechnology Co., Ltd, Shanghai, China). The RNA samples were diluted with RNA extract from nasopharyngeal and oropharyngeal swab of negative patients for detecting by RT-PCR.

#### Laboratory quality-control

Acceptable specimens are respiratory and serum specimens, the former including: nasopharyngeal and oropharyngeal swabs, bronchoalveolar lavage fluid, tracheal aspirates and sputum. Cotton swab heads are not allowed for swab specimens. Specimens should not be stored for more than 72 hours at 4 °C. Cross-reactive experiments should be performed to verify the specificity (Supplemental table 1). Positive control and negative control should be tested at the same time as all samples. The fluorescence amplification curve of negative control should not exceed the threshold. The CT value of all targets in the positive control should be within the expected range. The detection kit should contain the internal target gene, and the amplification curve should exceed the threshold line.

**Table 1.**
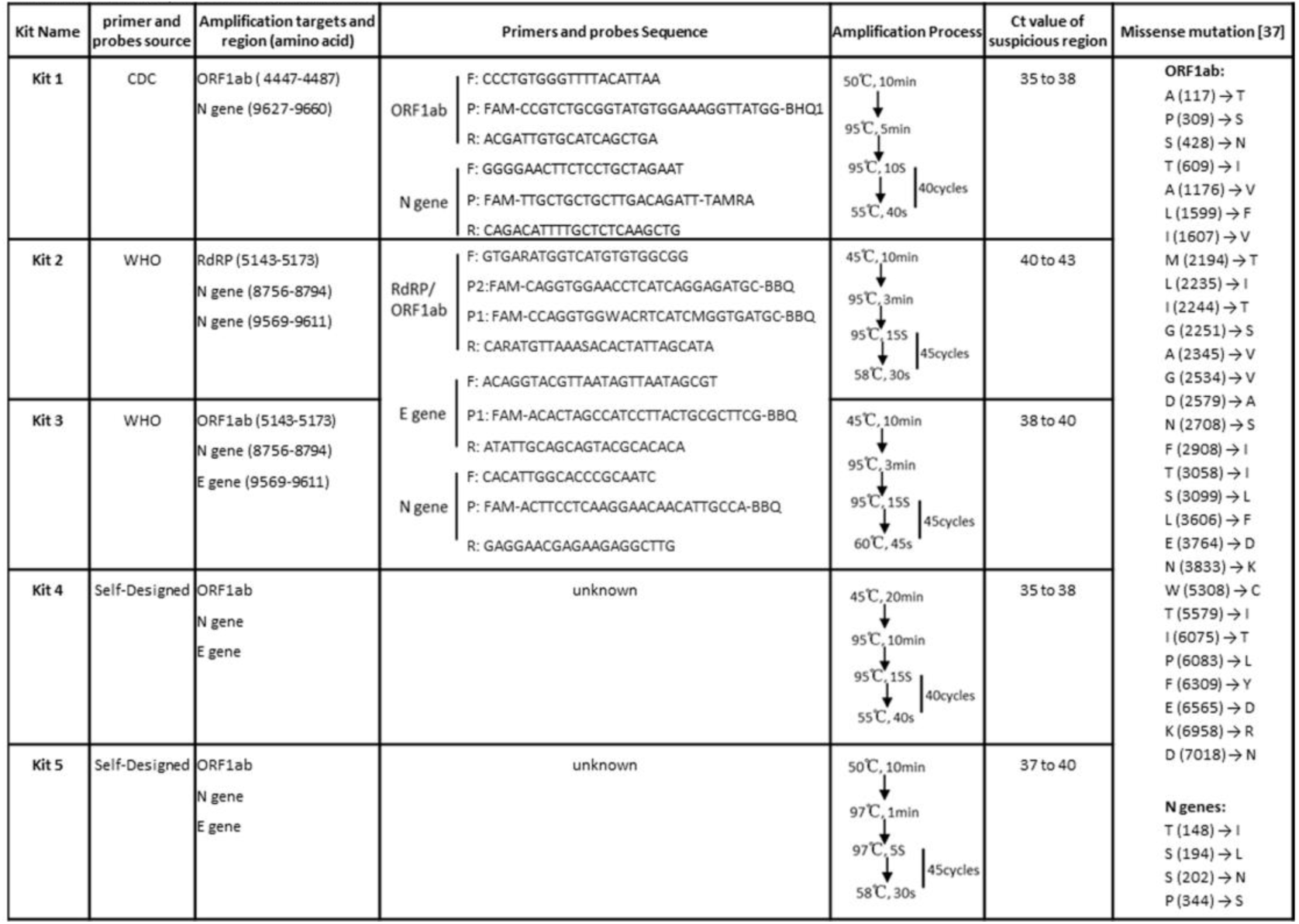
Information for the amplification Kits of SARS-CoV-2.

#### Real-time RT-PCR

Five different amplification kits were selected with three different primers and probes sources, among which one was from China’s CDC, two was from the World Health Organization (WHO) [6], and the other two were self-designed by the kit manufacturer. Information for the five amplification kits was shown in table 1. Each kit contained 25 μl of reaction system including 5μl of RNA template. The amplification was operated separately according to the instructions of kits. The amplification result was detected by ABI7500 Real-time PCR system (Applied Biosystems, USA).

#### Continuous amplification

The RT-PCR products were re-amplified for another 40 cycles under the same amplification conditions.

#### Statistical methods

SPSS18.0 software was used for statistical analysis. The Student *t* test was used to evaluate the differences between *Ct* values.

## RESULTS

### Sensitivity evaluation of SARS-CoV-2 detection kits

To verify the sensitivity of the kits, we took nasopharyngeal and oropharyngeal swab samples from a confirmed positive patient. After RNA extraction, the RNA was diluted according to the following proportion gradient: 1:5, 1:10, 1:20, 1:40, 1:80, 1:160, and 1:320. Then, RT-PCR results showed that the dilution titer of ORF1ab was the highest in the kit-1 (Figure 1A), indicating that the kit-1 was the most sensitive to SARS-CoV-2, followed by the kit-2. In addition, the CT value of the amplification curve was found to be positively correlated with the dilution titer (Figure 1B). The Ct values of ORF1ab gene and N gene in kit-1 were still within the reportable interval at 1:20 and 1:160 dilution respectively, while reached the detection suspicious region in kit-2 at 1:5 and 1:40 dilution titer.

**Figure 1.**
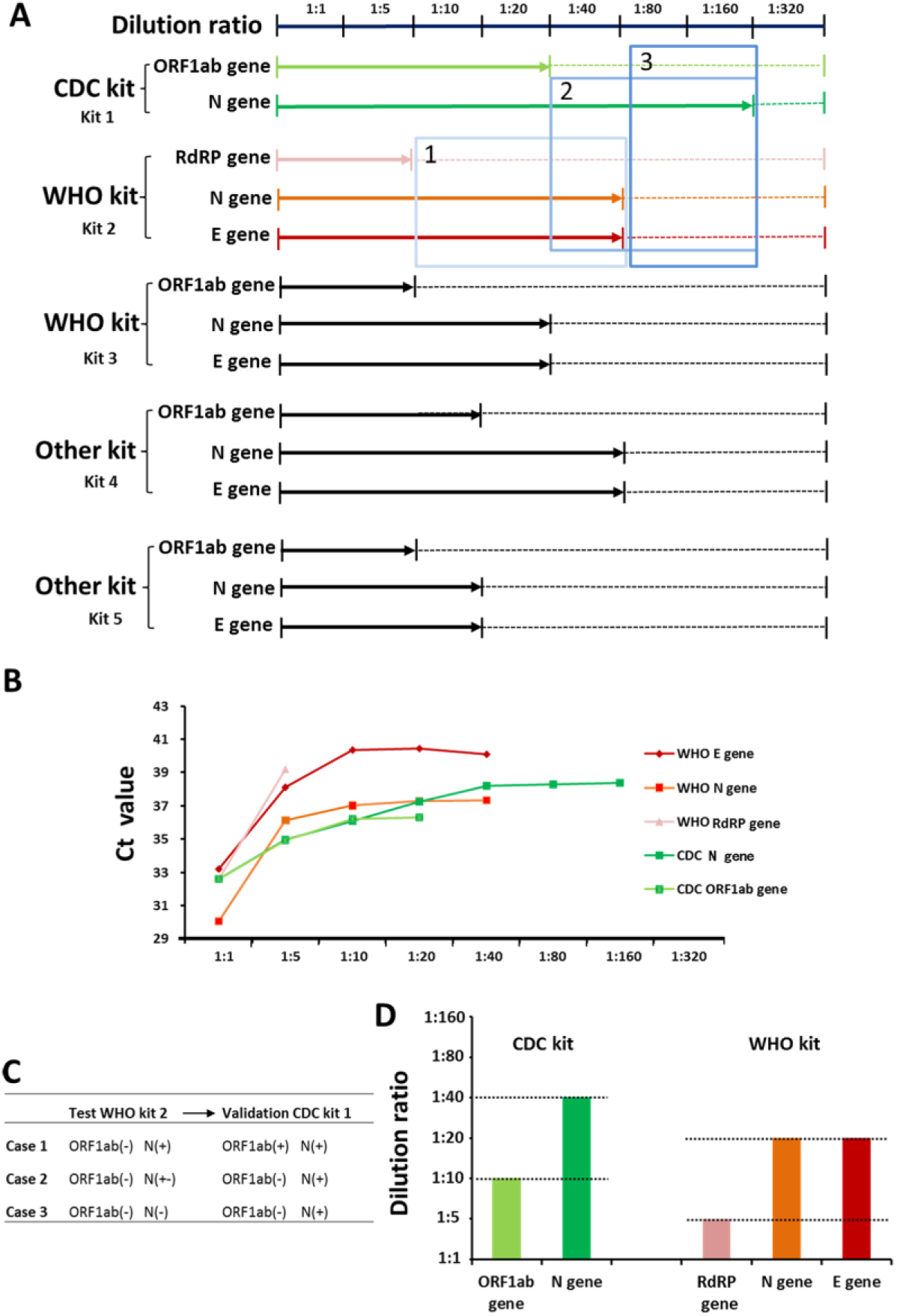
Sensitivity evaluation of SARS-CoV-2 detection kits. (A) Gradient dilution experiments showed that different kits have different sensitivity. (B) Ct value of different target genes were positively correlated with the dilution concentration. (C) Selection of test kit and validation kit. (D) CDC kit has the highest sensitivity through the verification of multiple positive samples.

Then, if the N gene or E gene is positive when ORF1ab gene is negative, how to judge the result and how to select the validation kit? For example, three cases were presented in in Figure.1A. Our solution is as follows (Figure. 1C): 1. Both ORF1ab and N genes can be converted to positive after verification with kit 1; 2. When N gene is in a suspicious region with kit 2, it can be converted to positive after verification with kit 1; 3. When N gene was negative with kit 2, it can be converted to positive after verification with kit 1. For the sake of further verify the sensitivity, another 9 positive samples were enrolled. The positive RNA extract was first quantified by digital PCR and then diluted to the same initial concentration. The results showed that ORF1ab gene can still be reported as positive at 1:10 dilution and the N gene even at 1:40 dilution (Figure. 1D) with kit-1, while they exceeded the detection line at 1:5 and 1:20 respectively with kit-2. Hence, we have reasons to believe that kit-1 has the highest sensitivity through the verification of multiple positive samples.

### Clinical validation and Application

Besides choosing a more sensitive kit for validation, is there an easier method to increase the positive detection rate? First, the RT-PCR products of the above diluted samples in the suspicious range were amplified for another 40 cycles, and found that for the samples with dilution gradients of 1:10 and 1:20, the ORF1ab and N genes with large original amplified Ct values were expanded to the positive reportable region, while other dilution gradients only with N or E genes were significantly amplified (figure. 2A). Moreover, 100 patients with clinical fever and dry cough who were suspected to be infected with the SARS-CoV-2 were enrolled for RT-PCR, and two positive cases and two suspicious cases were found (Table 2). Then, the suspicious cases were re-amplified to be positive by continuous amplification (Figure. 2B). Meanwhile, the environmental samples from 3 COVID-19 patients were conducted nucleic acid testing and found that the sample inside the mask of one patient was weakly positive, which could be reported as positive after another re-amplification (Figure 2C). Through analysis, we found that each target gene could reach the amplification plateau by adding another 30 cycles. In addition, we tried to add the initial RT-PCR amplification products of positive patients into a new amplification reaction system, and found that the results were not reliable (Data not shown).

**Figure 2.**
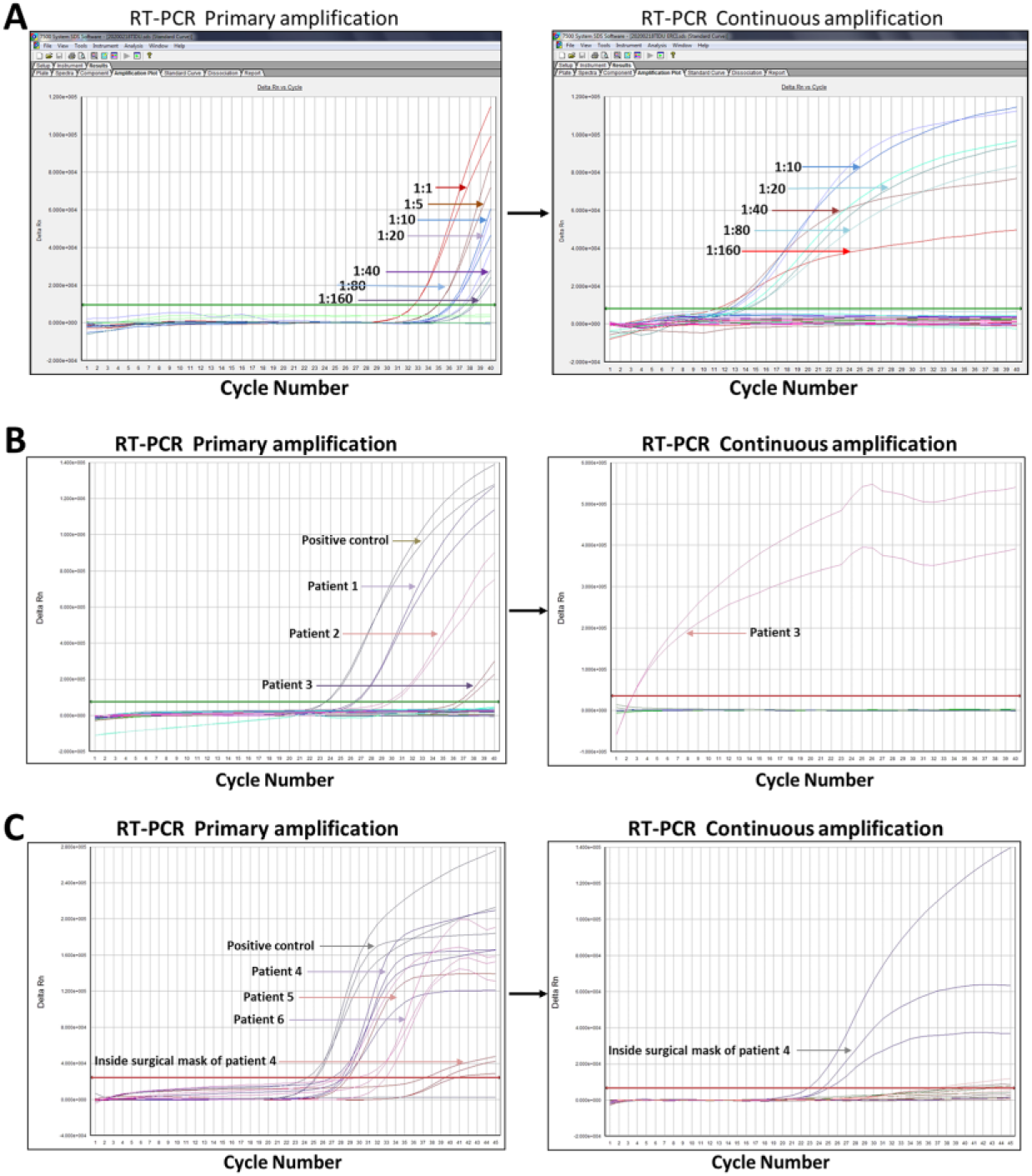
Strategies to reduce false negatives of SARS-CoV-2. (A) Continuous amplification of PCR products for gradient dilution samples. (B) Continuous amplification of PCR products for the nasopharyngeal and oropharyngeal swab specimens of clinical fever patients. (C) Continuous amplification of PCR products for the environmental samples of 3 positive patients. The specimen with a cycle threshold value of target genes above the baseline is interpreted as positive for SARS-CoV-2; those under, negative. Kit 1 was used for figure 2A and B, while kit 2 for figure 2C.

**Table 2.**
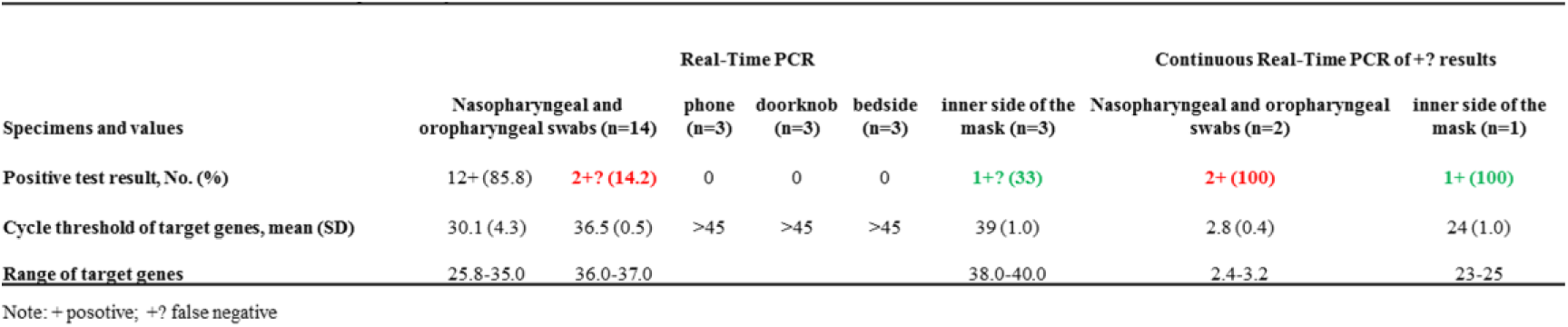
Detection Results of Clinical environmental specimens by Real-Time PCR.

### Strategies to reduce false negatives of SARS-CoV-2

Above all, we suggest that the laboratory must evaluate the sensitivity of detection kits first. If the laboratory can only select two kits, the selection strategy from detection to validation kit should be based on its sensitivity from low to high. In other words, the sensitivity of the validation kit must be higher than that of the test kit, which is of great significance to the suspected re-examination samples and the discharge criteria of patients. Moreover, for these specimens with the suspicious interval region or single channel positive results, the continuous amplification can be used to increase the detection rate of low viral load specimens and greatly reduce the false negative rate of SARS-CoV-2.

## DISCUSSION

As of 28^th^ April 2020, statistical data showed that the global number of confirmed cases of COVID-19 had surpassed 3000,000 with more than 210,000 deaths. With an increasing number of potential cases emerge, the SARS-CoV-2 poses a major threat to global public health[34]. A greater number of diagnostic tools have been developed such as virus isolation, PCR-based assays, IHC, and antibody assays, which are currently in place across different diagnostic laboratories around the world[35, 36]. Although, RT-PCR is challenged by the “false negative” results[37], in view of the past major epidemic outbreaks [38], RT-PCR is still the preferred detection method, and how to ensure the accuracy of nucleic acid test results is the currently facing problem. In this study, we aimed to evaluate the sensitivity of different RT-PCR kits for COVID-19 diagnosis.

Different target genes of SARS-CoV-2 were used in countries (https://www.who.int/emergencies/diseases/novel-coronavirus-2019/technical-guidance/laboratory-guidance), while some scholars believe that the detection of N gene is an accurate, rapid, early and simple diagnostic method for COVID-19[39]._Then, under the current circumstance, can the single target N or double targets N and E be judged as positive for SARS-CoV-2? An increasing number of articles showed that the SARS-CoV-2 is undergoing rapid mutation [40, 41]. Ninety-three mutations were found over the entire genomes of SARS-CoV-2 [42] with twenty-nine missense mutations in the ORF1ab region and four in the N region (table1). Fortunately, through gene comparison on BLAST, the primer or probe sequences published by CDC and WHO were not in these mutation regions. Moreover, study found that a deletion of 382 nucleotides in the ORF8 gene can enhances the transcription of the downstream N gene[28] which may increase the false negative detection rate of SARS-CoV-2. Thus, attentions should be paid to the abnormally amplified N gene in clinical detection. The latest clinical research has revealed that the prevalence of SARS-CoV-2 among patients with influenza like illnesses reached 5% in Los Angeles medical center[43]. Due to the difficulty to distinguish the mild COVID-19 with influenza virus [44–46], China CDC has made it clearly that influenza testing should be combined with SARS-CoV-2 testing [47].

Although the detection rate of viral nucleic acid is closely related to the course of viral infection, which is not completely clear and the optimal sampling time is uncertain, so it is likely that the period of high viral load in the nasopharyngeal swab samples will be missed, resulting in false negatives[48]. The nasopharyngeal swabs are the most common samples for SARS-CoV-2 detection, while the stool samples are reported as a good test target during treatment recently[49], and the positive fecal nucleic acid has an important correlation with the development of the patient’s disease course, which may become an important indicator for the evaluation of therapeutic efficacy and discharge criteria.

As with all viral nucleic acid testing projects, the RT-PCR results of SARS-CoV-2 are affected by various factors including before, during and after detection, thus sufficient laboratory quality-control measures should be taken. Although, the continuous amplification and other detection methods of SARS-CoV do exhibit false positive results [50, 51], the continuous amplification is recommend to use only when the amplification curve of target gene is in the specious region. More importantly, we also believe that antibody test and nucleic acid test should complement each other to improve the diagnosis effect, especially to screen asymptomatic patients better, so as to reduce the detection “false negative” phenomenon of “false recovered patients” or premorbid patients with low virus latency.

### Declarations

#### Ethics approval and consent to participate

This study was undertaken with the approval of the Jinan Central Hospital Affiliated to Shandong University Ethics Service Committee. All experiments were carried out in accordance with the approved guidelines. Informed consent was obtained from all the participants prior to sampling.

## Data Availability

yes

## Acknowledgements

We thank members of our laboratories for helpful discussions.

## Authors’ contributions

YW conceived and designed the study. YZ was the major contributor in drafting the manuscript. YZ and QZ performed the experiments. LW and HZ collected the samples. FP, HL, MJ, WY and QW extracted the RNA and made the clinical diagnoses. All authors read and approved the final manuscript.

